# Detection and whole-genome sequencing of SARS-CoV-2 B.1.617.2 and B.1.351 variants of concern from Pakistan during the COVID-19 third wave

**DOI:** 10.1101/2021.07.14.21259909

**Authors:** Massab Umair, Aamer Ikram, Muhammad Salman, Nazish Badar, Syed Adnan Haider, Zaira Rehman, Muhammad Ammar, Muhammad Suleman Rana, Qasim Ali

## Abstract

The viral lineages reflecting variants of concern have emerged worldwide and among them B.1.1.7 (Alpha), B.1351 (Beta) and B.1.617.2 (Delta) variants are the most significant ones and merit close monitoring. In Pakistan, very limited information is available on the circulation of these variants and only the alpha variant has been reported as the main circulating lineage. The objective of this study was to detect and explore the genomic diversity of B.1.351 and B.1.617.2 during the third wave in the indigenous population. During the current study, a total of 2274 samples were tested on real-time PCR for the presence of SARS-CoV-2 from May 14 to May 31, 2021, and among these, 17% were spike positive, whereas 83% of samples had the spike gene target failure (SGTF). From these spike positive samples, 22 samples were processed for whole-genome sequencing. Among VOCs, 45.5% (n=10) belonged to B.1.351 followed by B.1.617.2, 36% (n=8). The delta variant cases were reported mostly from Islamabad (n = 5; 63%) followed by Peshawar and Azad Kashmir (n = 1; 13% each). Beta variant cases originated from Islamabad (n=5; 56%), Peshawar (n=2; 22%), Azad Kashmir and Rawalpindi (n=1; 11% each). The mutation profile of delta variant isolates reported significant mutations, L452R, T478K, P681R, and D950N. The beta variant isolates reported characteristic mutations, D215G, K417N, E484K, N501Y, and A701V. Notably, a characteristic mutation, E484Q was also found in delta variant, B.1.617.2. Our current findings suggest detection of these VOCs from the local population and warrants large-scale genomic surveillance in the country. In addition, it provides timely information to the health authorities to take appropriate actions.

## INTRODUCTION

The COVID-19 Pandemic is still one of the leading causes of infectious mortality worldwide in the advent of the emergence of novel SARS-CoV-2 variants. As of June 20, 2021, the global total of SARS-CoV-2-related infections has surpassed over 179.1 million, with 3.86 million deaths. In recent months, a diversification of SARS-CoV-2 has been observed globally as a result of evolution and adaptation processes. Some emerging mutations may confer a selective advantage to the virus, resulting in the selection of the “variants of concern” (VOCs) with significant epidemiological and pathogenic consequences [1, 2]. Four specific viral lineages reflecting variants of concern have emerged worldwide and warrant close monitoring: B.1.1.7 (alpha), B.1.351 (beta), P.1 (gamma), and B.1.617.2 (delta) [3]. Among them, alpha, beta, and delta variant has been recently the most important Variant of Concern which has contributed significantly to the upsurge of new waves worldwide.

The beta variant, B.1.351, is reported in about 95 countries including Pakistan as of June 20, 2021, and is characterized by seven different lineage defining variants in the spike protein region, with three significant mutations, N501Y, E484K and K417N at particular residue sites called receptor-binding domain (RBD) [4]. There is evidence that South African strains have increased transmissibility as a result of the N501Y substitution [5] due to the increased binding affinity of human angiotensin-converting enzyme 2 (ACE2) in a mouse model, as determined by deep mutation scanning [6, 7]. Additionally, the South African variant may provide an immune escape mechanism from antibodies due to the E484K mutation in the spike protein [8]. It was also recently identified as a variant capable of evading monoclonal and serum antibody responses [9, 10].

The variant B.1.617 originated in Maharashtra and has since spread throughout the state and to a number of other countries. The sub-lineage, B.1.617.1 is defined by the presence of constellation of mutations, L452R, P681R and E484Q in spike region, whereas B.1.617.2 is characterized by the spike mutations, L452R, P681R and T478K. The RBD mutations enhance infectivity due to presence of L452R and T478K by increasing the spike protein’s affinity for the human ACE2 receptor [11, 12]. Both mutations reduce the binding affinity of monoclonal antibodies, thereby impairing their neutralizing ability. Additionally, structural analysis of mutations in the furin cleavage site, L452R, and E484Q, as well as P681R, revealed increased ACE2 binding and cleavage rate, resulting in increased transmissibility [13]. Keeping in view the emergence of B.1.351 and B.1.617.2 variants of concern, we aimed to detect and explore the genomic diversity of these VOCs in Pakistan.

## MATERIALS AND METHODS

### Sampling

Oropharyngeal samples (n = 2274) were collected from the COVID-19 suspected patients received at the National Institute of Health’s Department of Virology in Islamabad. As the spike gene target failure (SGTF) was used as a proxy for the detection of the B.1.1.7 variant, on TaqPath™ kit (ThermoFisher Scientific, Waltham, US) [13]. This detection method can also be used the other way around for surveillance of lineages other than B.1.1.7. Based on this strategy, for whole-genome sequencing, we initially screened spike positive samples having low cycle threshold (Ct) values (< 27 of S gene) followed by selection based on their geographical location i.e., representing the whole country.

### RNA Extraction, cDNA synthesis, and Amplification

The viral RNA was isolated from patient samples according to the standard protocol using a QIAamp Viral RNA Mini Kit (Qiagen, Germany). The cDNA synthesis and amplification were performed according to the Primal-Seq Nextera XT protocol (version 2) using SuperScript™ IV VILO™ Master Mix (Invitrogen, USA) and Q5® High-Fidelity 2X Master Mix (New England BioLabs, USA), with the ARTIC nCoV-2019 Panel (Integrated DNA Technologies, Inc, USA) [14].

### Next Generation Sequencing

The paired-end sequencing library (2×150 bp) was prepared from the generated amplicons using the Illumina DNA Prep Kit (Illumina, Inc, USA) by following the standard protocol. The prepared libraries were pooled and subjected to sequencing on Illumina platform, iSeq using sequencing reagent, iSeq 100 i1 Reagent v2 (300-cycle) (Illumina, Inc, USA) at Department of Virology, National Institute of Health, Islamabad, Pakistan.

### Data Analysis

The FastQC tool (v0.11.9) was used to assess the read quality of sequenced files [16]. Trimmomatic (v0.39) [15] was employed to remove Illumina adapter sequences and low-quality base calls with scores less than 30 to eliminate technical biases and artifacts. The filtered reads were assembled by aligning with the available reference genome of SARS-CoV-2 (Accession number: NC_045512.2) using the Burrows-Wheeler Aligner’s (BWA, v0.7.17) default settings [16]. Picard Tools was used to remove PCR duplicates from aligned reads (v2.25.4) [17]. The variants were called and consensus sequences of all genomes were generated as per Centers for Diseases Control and Prevention (CDC, USA) guidelines [18]. The assembled genomes were classified into PANGO lineages using the Pangolin v3.1.5 and pangoLEARN model dated 15-06-2021 [19].

### Phylogenetic Analysis

Nextstrain’s standard protocol for analyzing SARS-CoV-2 genomes was used for the phylogenetic analysis. To begin, BLAST searches were conducted on the GISAID database against all SARS-CoV-2 genome sequences for each of the current study’s beta and delta isolates. This resulted in a total of 78 sequences including the current study’s sequences. The sequences were clustered using Augur, Nextstrain’s phylodynamic pipeline [20]. Alignment of the sequences to the Wuhan reference genome was performed using MAFFT v7.470 [21]. The initial phylogenetic tree was constructed using IQTREE v1.6.12 [22], which implements the Augur tree using the generalized time reversible (GTR) model and performs bootstrapping to ensure a high degree of confidence in the tree topology. Using the Wuhan reference genome (GISAID ID: EPI_ISL_406798), the raw tree was rooted. TreeTime v0.8.1 was used to further process the tree to generate a time-resolved phylogeny based on maximum likelihood [23]. The resulting tree was visualized using Auspice.

## RESULTS

From May 14 to May 31, 2021, a total of 2274 samples were tested using the TaqPath™ COVID-19 kit (ThermoFisher Scientific, US) for the presence of SARS-CoV-2. Among these, 17% (n = 377) of samples exhibited spike gene amplification, whereas 83% (n = 1897) of samples exhibited spike gene target failure (SGTF) (Figure 1). From these 377 spike positive samples, 22 samples were selected for whole-genome sequencing based on the criteria defined above.

**Figure 1:**
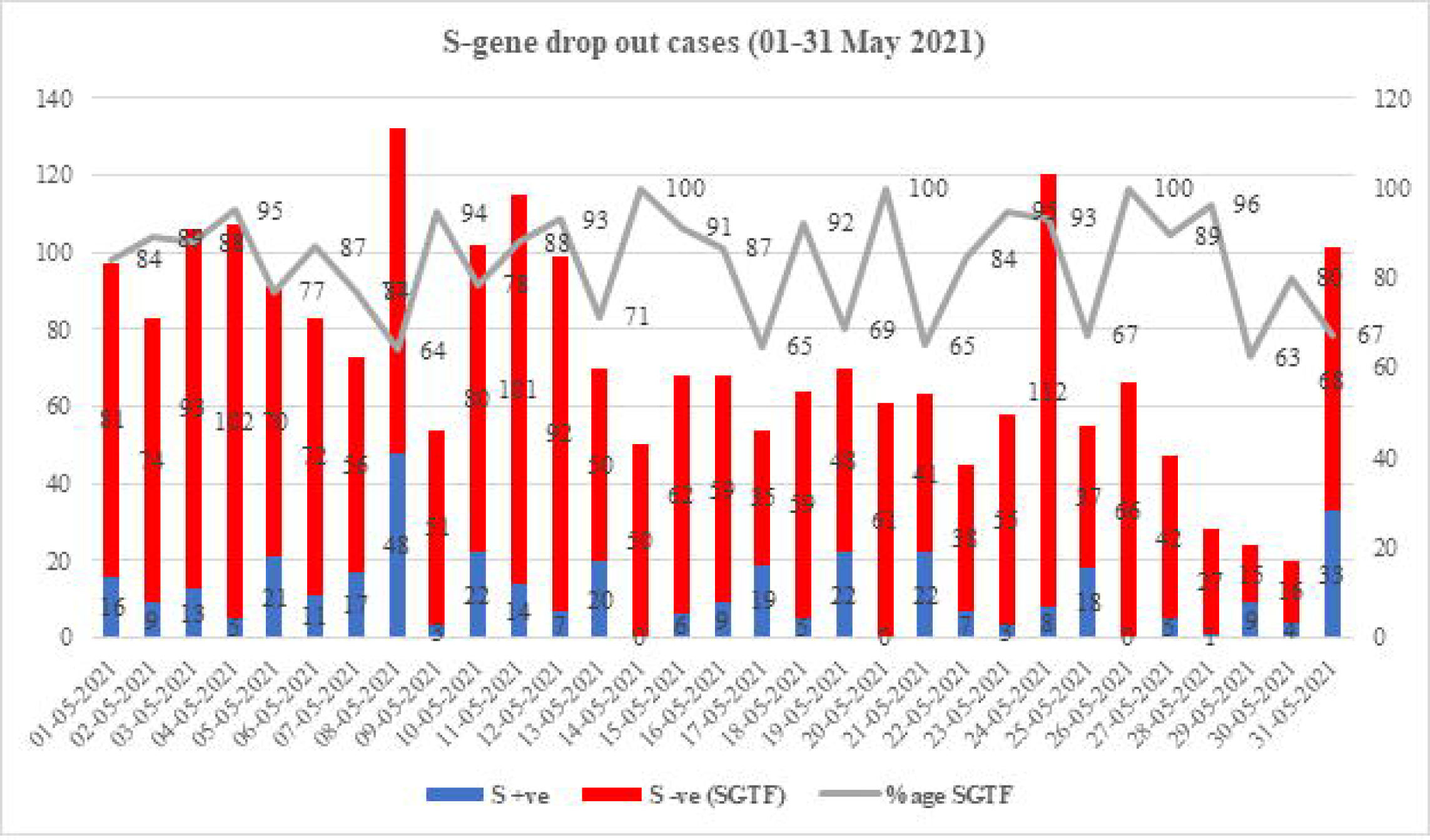
The distribution of SARS-CoV-2 cases according to the Δ69–70 deletion during May 2021. SGTF was detected in SARS-CoV-2 patients using the TaqPath real-time PCR kit. The X-axis indicates the months, while the Y-axis indicates the number of cases. S+ve indicates the presence of a spike gene; SGTF indicates the absence of a spike gene.

In the present study, a total of six lineages were identified including the delta VOC B.1.617.2 for the first time in Pakistan. Among VOCs, 45.5% (n=10) belonged to B.1.351 followed by B.1.617.2, 36% (n=8). The remaining, 18.5% (n=4), reported lineages are B.1.1.448, B.1.36, and B.1. The delta variant cases were reported mostly from Islamabad (n = 5; 63%) followed by Peshawar and Azad Kashmir (n = 1; 13% each). Beta variant cases originated from Islamabad (n=5; 56%), Peshawar (n=2; 22%), Azad Kashmir and Rawalpindi (n=1; 11% each) (Figure 2). Two patients infected with delta variant had been associated with international travel with one traveled back from Saudi Arabia (GISAID ID: EPI_ISL_2434174) and the other from Middle East (GISAID: EPI_ISL_2438666). Furthermore, the travel history of two male patients from the Middle East with beta variant was also reported (GISAID ID: EPI_ISL_2438665 and EPI_ISL_2438683)

**Figure 2:**
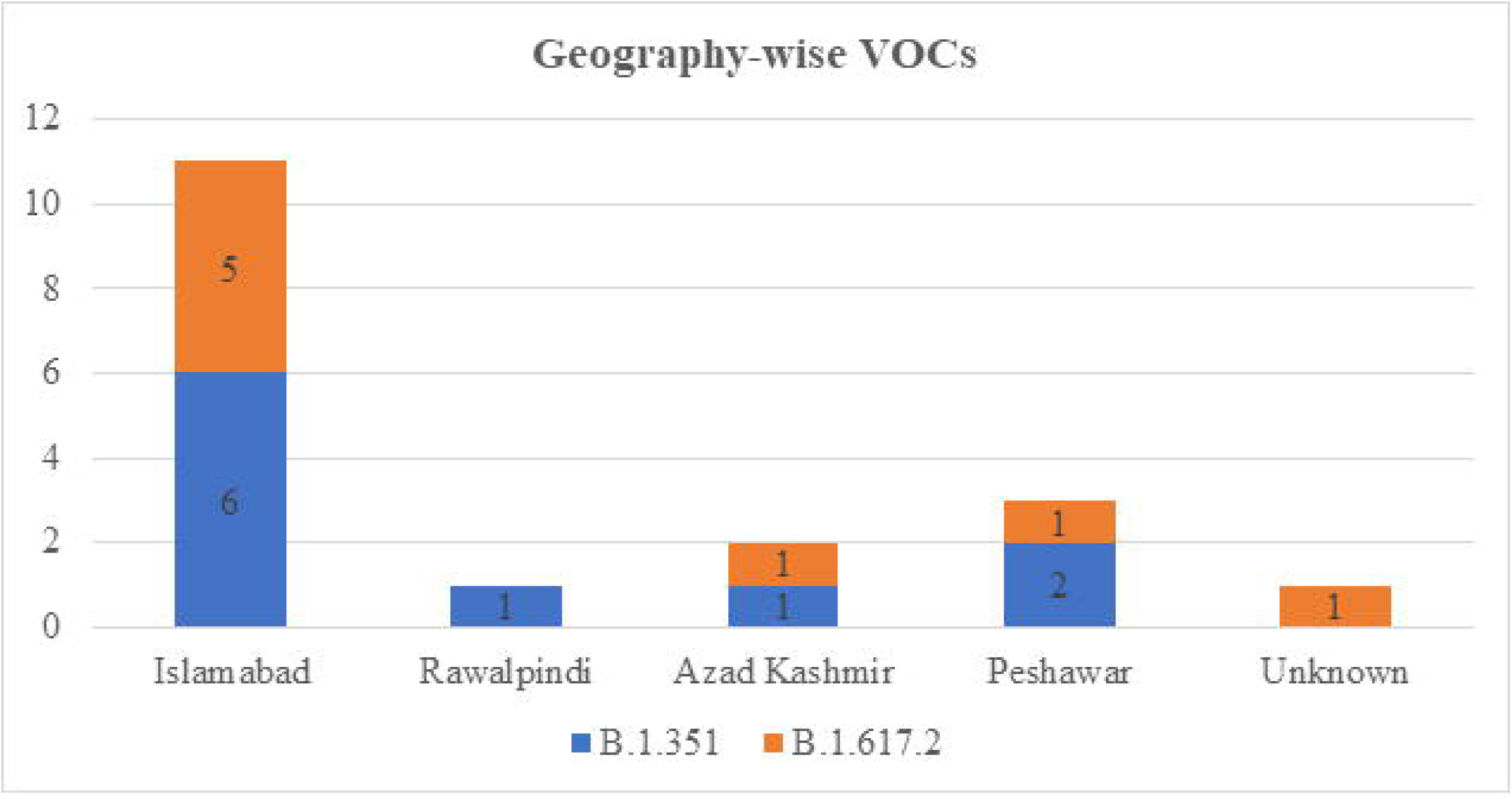
Geography-wise distribution of variants of concerns reported in the current study. X-axis representing the geographical location and the Y-axis representing the number of delta B.1.617.2 and beta B.1.351 and sequences.

Patients infected with the beta variant had a female to male infection ratio of 1:1, with 50% of females and 50% of males infected. However, the female to male infection ratio in case of delta was 3:5, with 37.5 percent females and 62.5 percent males infected (Figure 3).

**Figure 3:**
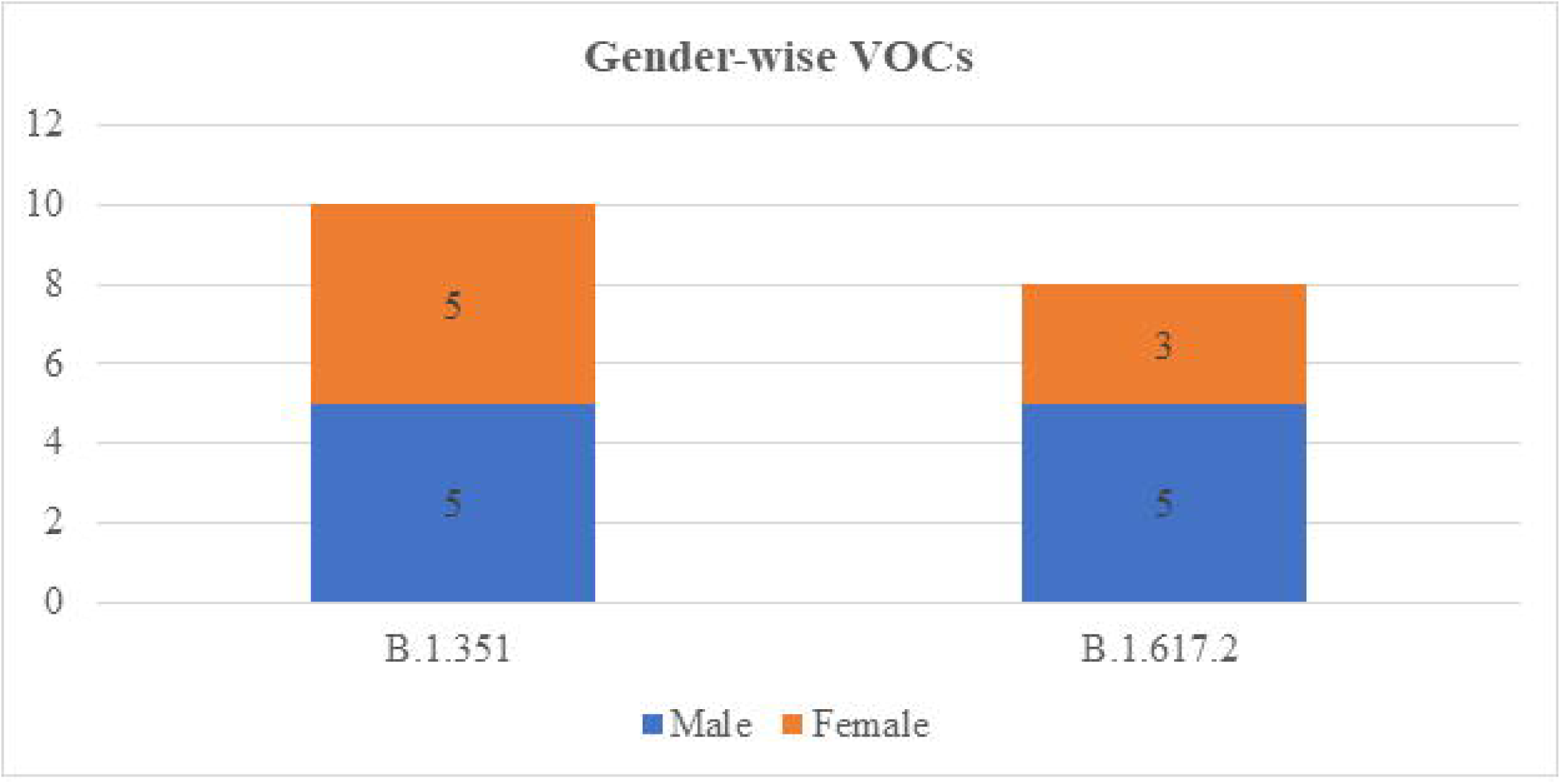
Gender-wise distribution of variants of concerns reported in the current study. X-axis representing the delta B.1.617.2 and beta B.1.351 variant and Y-axis representing the number of sequences associated with gender.

The mutation profile of delta variant, B.1.617.2, isolates reported significant mutations, S:L452R (22917T>G), S:T478K (22995C>A), S:P681R (23604C>G), S:D950N (24410G>A), ORF3a:S26L (25469C>T), M:I82T (26767T>C), ORF7a:V82A (27638T>C), ORF7a:T120I (27752C>T), N:D63G (28461A>G), N:R203M (28881G>T) and N:D377Y (29402G>T). A highly significant mutation, E484Q (23012G>C), having a role in the immune escape was also found in a male patient (GISAID ID: EPI_ISL_2434972).

The mutational analysis revealed that all the beta variant isolates reported significant and lineage defining mutations, ORF1a:K1655N (5230G>T), S:D80A (21801A>C), S:D215G (22206A>G), S:K417N (22813G>T), S:E484K (23012G>A), S:N501Y (23063A>T), S:A701V (23664C>T), E:P71L (26456C>T) and N:T205I (28887C>T). Corresponding to the person who traveled back to Pakistan from the Middle East reported several unique missense mutations in spike region, A27S (21641G>T), G181V (22104G>T), ORF1b region, L820F (15925C>T), ORF3a region, T271I (26204C>T) and N region, K387N (29434G>T) (GISAID ID: EPI_ISL_2438683).

Another Middle East traveler, GISAID ID: EPI_ISL_2438665, carried one unique mutation in the N region (T362I (29358C>T). Three distinct mutations in ORF1a region, T183I (813C>T), S3099L (9561C>T) and L3915F (12008C>T) were observed in one of the isolates (GISAID ID: EPI_ISL_2313081). Furthermore, distinct spike mutations, A879S (24197G>T) and D1163Y (25049G>T) were also found in a patient with unknown travel history (EPI_ISL_2313084). The detailed information about the infected patients with VOCs along with their distinct amino acid mutations is shown in Table 1.

**Table 1:** Details of the patients infected with VOCs along with the amino acid mutations found in the study enrolled samples.

| Sr# | Lab ID (NIH) | GISAID ID | Gender | Location | Travel History | Lineage | Distinct Amino Acid Mutations |
| --- | --- | --- | --- | --- | --- | --- | --- |
| 1 | NIH-B10-S7 | EPI_ISL_2313081 | Male | Islamabad | Not available | B.1.351 | ORF1a:K1655N, S:D80A, S:D215G, S:K417N, S:E484K, S:N501Y, S:A701V, E:P71L and N:T205I |
| 2 | NIH-B10-S11 | EPI_ISL_2313084 | Female | Islamabad | Not available | B.1.351 | ORF1a:K1655N, ORF1a: S911F, ORF1b: T1511I, S:D80A, S:D215G, S: A879S, S:K417N, S: D1163Y, S:E484K, S:N501Y, S:A701V, E:P71L and N:T205I |
| 3 | NIH-B10-S12 | EPI_ISL_2313086 | Male | Islamabad | Not available | B.1.617.2 | S:T19R, S:L452R, S:T478K, S:P681R, S:D950N, ORF3a:S26L, M:I82T, ORF7a:V82A, ORF7a:T120I, N:D63G, N:R203M and N:D377Y |
| 4 | NIH-B10-S16 | EPI_ISL_2313098 | Female | Islamabad | Not available | B.1.351 | ORF1a:K1655N, S:D80A, S:D215G, S:K417N, S:E484K, S:N501Y, S:A701V, E:P71L and N:T205I |
| 5 | NIH-B10-S17 | EPI_ISL_2313099 | Female | Islamabad | Not available | B.1.351 | ORF1a:K1655N, S:D80A, S:D215G, S:K417N, S:E484K, S:N501Y, S:A701V, E:P71L and N:T205I |
| 6 | NIH-B10-S18 | EPI_ISL_2313112 | Female | Islamabad | Not available | B.1.351 | ORF1a:K1655N, S:D80A, S:D215G, S:K417N, S:E484K, S:N501Y, S:A701V, E:P71L and N:T205I |
| 7 | NIH-B10-S19 | EPI_ISL_2313114 | Male | Islamabad | Not available | B.1.351 | ORF1a:K1655N, S:D80A, S:D215G, S:K417N, S:E484K, S:N501Y, S:A701V, E:P71L and N:T205I |
| 8 | NIH-B10-S20 | EPI_ISL_2314185 | Male | Rawalpindi | Not available | B.1.351 | ORF1a:K1655N, S:D80A, S:D215G, S:K417N, S:E484K, S:N501Y, S:A701V, E:P71L and N:T205I |
| 9 | NIH-B11-S1 | EPI_ISL_2434174 | Female | Islamabad | Saudi Arabia | B.1.617.2 | S:T19R, S:L452R, S:T478K, S:P681R, S:D950N, ORF3a:S26L, M:I82T, ORF7a:V82A, ORF7a:T120I, N:D63G, N:R203M and N:D377Y |
| 10 | NIH-B11-S2 | EPI_ISL_2434247 | Female | Azad Kashmir | Not available | B.1.351 | ORF1a:K1655N, ORF1a: T183I, ORF1a: S3099L, ORF1a: L3915F S:D80A, S:D215G, S:K417N, S:E484K, S:N501Y, S:A701V, E:P71L and N:T205I |
| 11 | NIH-B11-S3 | EPI_ISL_2434781 | Male | Islamabad | None | B.1.617.2 | S:T19R, S:L452R, S:T478K, S:P681R, S:D950N, ORF3a:S26L, M:I82T, ORF7a:V82A, ORF7a:T120I, N:D63G, N:R203M and N:D377Y |
| 12 | NIH-B11-S4 | EPI_ISL_2434972 | Male | Islamabad | Bahrain | B.1.617.2 | S:T19R, S:L452R, S:T478K, S:P681R, S:D950N, ORF3a:S26L, M:I82T, ORF7a:V82A, ORF7a:T120I, N:D63G, N:R203M and N:D377Y |
| 13 | NIH-B11-S5 | EPI_ISL_2434976 | Female | Islamabad | None | B.1.617.2 | S:T19R, S:L452R, S:T478K, S:P681R, S:D950N, ORF3a:S26L, M:I82T, ORF7a:V82A, ORF7a:T120I, N:D63G, N:R203M and N:D377Y |
| 14 | NIH-B11-S7 | EPI_ISL_2434982 | Female | Not available | Not available | B.1.617.2 | S:T19R, S:L452R, S:T478K, S:P681R, S:D950N, ORF3a:S26L, M:I82T, ORF7a:V82A, ORF7a:T120I, N:D63G, N:R203M and N:D377Y |
| 15 | NIH- B11-S8 | EPI_ISL_2438547 | Male | Azad Kashmir | Not available | B.1.617.2 | S:T19R, S:L452R, S:T478K, S:P681R, S:D950N, ORF3a:S26L, M:I82T, ORF7a:V82A, ORF7a:T120I, N:D63G, N:R203M and N:D377Y |
| 16 | NIH- B11-S13 | EPI_ISL_2438665 | Male | Peshawar | Middle East | B.1.351 | ORF1a:K1655N, S:D80A, S:D215G, S:K417N, S:E484K, S:N501Y, S:A701V, E:P71L, N: T362I and N:T205I |
| 17 | NIH-B11-S14 | EPI_ISL_2438666 | Male | Peshawar | Middle East | B.1.617.2 | S:T19R, S:L452R, S:T478K, S:P681R, S:D950N, ORF3a:S26L, M:I82T, ORF7a:V82A, ORF7a:T120I, N:D63G, N:R203M and N:D377Y |
| 18 | NIH-B11-S15 | EPI_ISL_2438683 | Male | Peshawar | Middle East | B.1.351 | ORF1a:K1655N, ORF1b: L820F, ORF3a: T271I S:D80A, S: A27S, S:D215G, S: G181V, S:K417N, S:E484K, S:N501Y, S:A701V, E:P71L N:T205I and N:K387N |

To determine the origin of Pakistani isolates, we constructed a maximum likelihood phylogenetic tree using 79 SARS-CoV-2 virus full-length genomes. In the phylogenetic tree, eighteen viruses from the Pakistani population were dispersed across 2 SARS-CoV-2 clades, GH (Beta) and G (Delta) (Figure 4) depicting close similarities with isolates originating from Asia, Europe, and North America. The beta-variant infected patients (GISAID ID: EPI_ISL_2438665 and EPI_ISL_2438683) having a travel history of the Middle East, clustered mostly with USA, India, and England viral isolates (Figure 5). The close similarities of one delta variant patient (travel history of Saudi Arabia) were observed mostly with Bangladesh followed by India and England viral isolates (GISAID ID: EPI_ISL_2434174). Also, another case (GISAID: EPI_ISL_2438666) with a Middle East travel history showed close association with Indian isolates (Figure 6). This result suggests the introduction of VOCs through the importation of cases from different countries.

**Figure 4:**
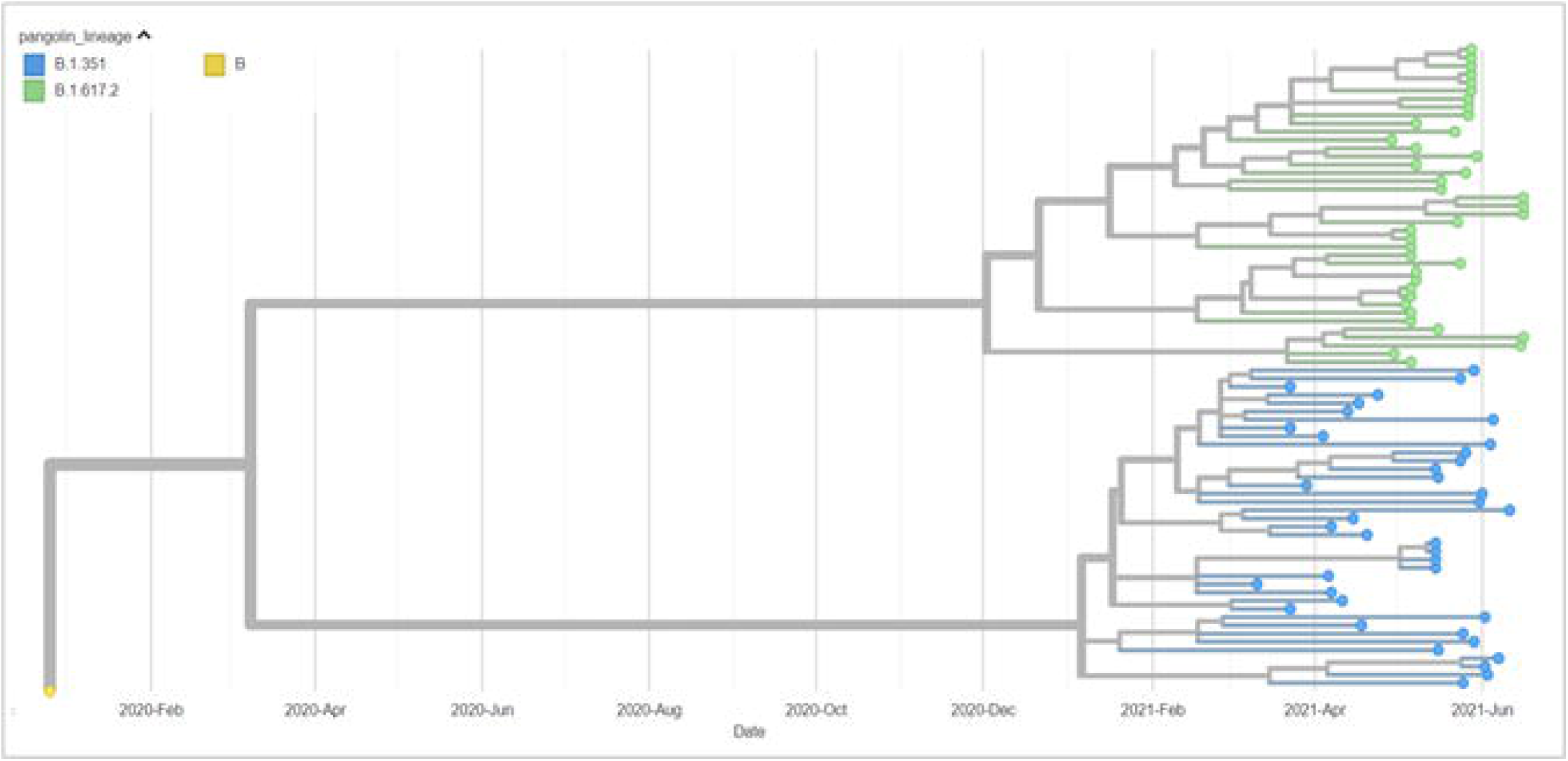
Phylogenetic distribution of beta and delta lineages in 78 SARS-CoV-2 viral genomes from around the world, including current study sequences from Pakistan with reference to the Wuhan reference genome (EPI ISL 402125). The maximum likelihood phylogenetic tree was constructed using Nextstrain’s Augur tree pipeline, and IQ-TREE was run with the default parameters. The time-resolved phylogenetic tree with selected metadata information was constructed using TreeTime and visualized in Auspice.

**Figure 5:**
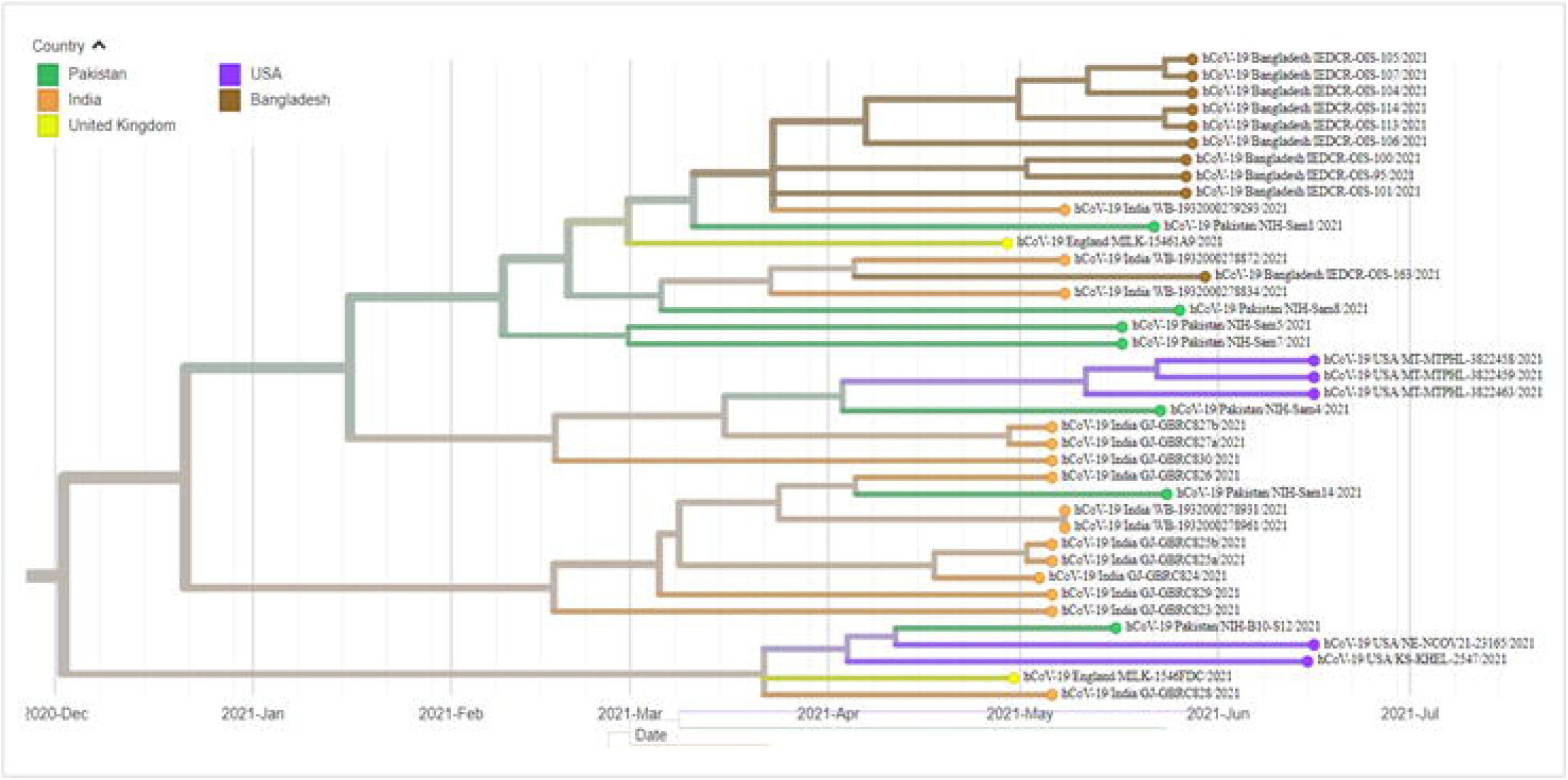
Phylogenetic worldwide distribution of delta B.1.617.2 lineages from 39/78 SARS-CoV-2 viral genomes including current study sequences from Pakistan.

**Figure 6:**
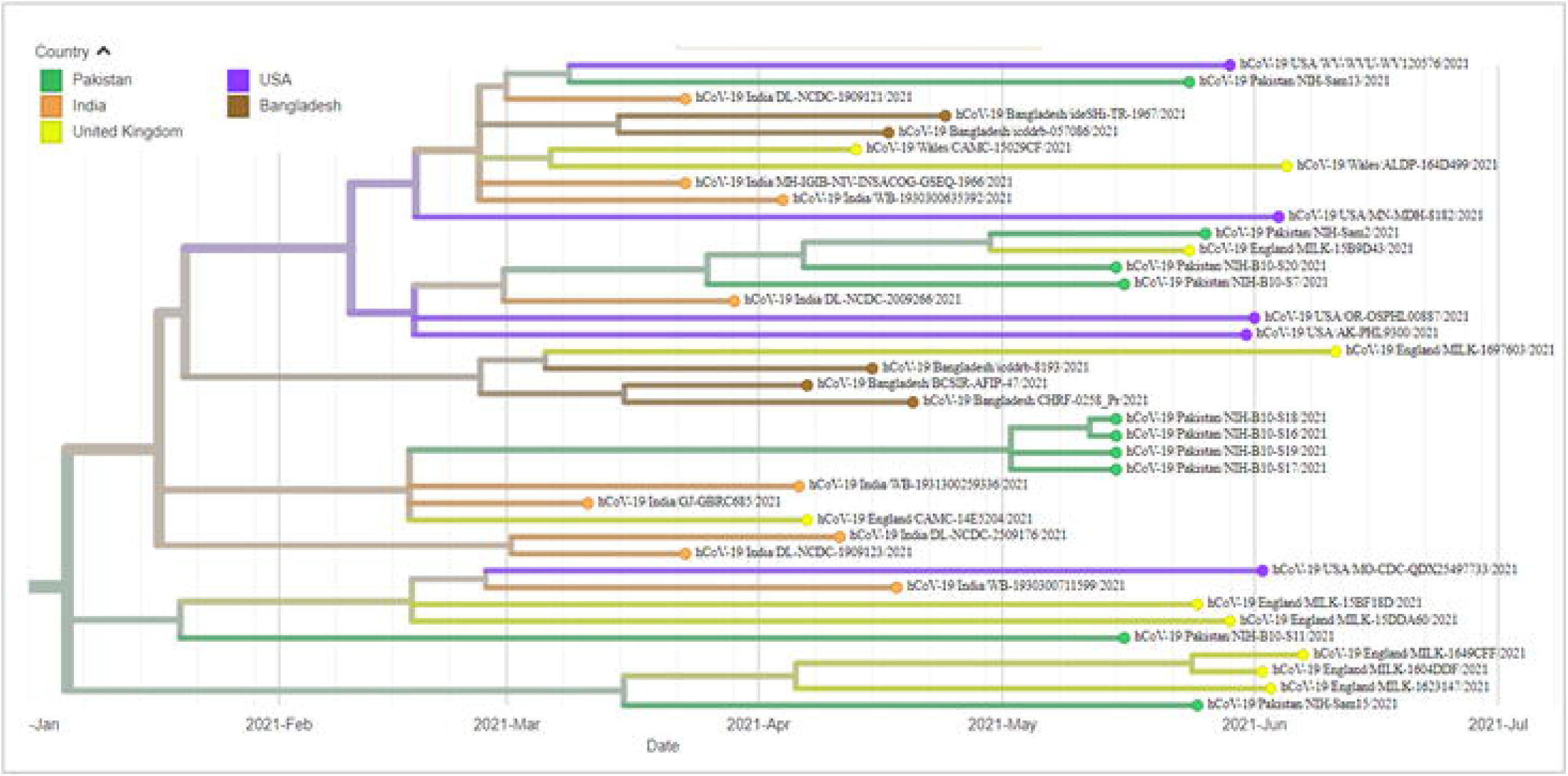
Phylogenetic worldwide distribution of beta B.1.317 lineages from 39/78 SARS-CoV-2 viral genomes including current study sequences from Pakistan.

## DISCUSSION

The COVID-19 pandemic has affected every continent, resulting in a global health crisis that has been aggravated by the recent emergence of different variants of the virus. Amongst the SARS-CoV-2 variants of concern, B.1.1.7 (Alpha) became the predominant lineage worldwide after the initial detection from the UK in September 2020. The first case of B.1.1.7 from Pakistan was reported in December 2020 [24] followed by a rapid spread of this infectious variant in the indigenous population [25] that triggered a spike in cases in March 2021 leading to the third wave. Although the prevalence and sustained circulation of B.1.1.7 in Pakistan is well established, data on the presence and genomic diversity of other lineages and VOCs is limited mainly due to the lack of genomic surveillance. Therefore, the aim of the current study was to detect and explore the genomic diversity of SARS-CoV-2 variants (other than B.1.1.7) prevalent in Pakistan during the third wave. Our results showed a high percentage of B.1.617.2 (Delta) (n=8) and B.1.351 (Beta) (n=10) variants among the samples selected for whole-genome sequencing during May-June 2021 and serve as the first report on the detection and exploration of the genomic diversity of delta variant from Pakistan. The delta variant, which was first identified from India during September 2020, caused a ferocious second wave in the country with 414,188 cases reported on 06 May 2021. As of 24 June 2021, the variant has been detected from 77 countries and linked to the recent surge in COVID-19 cases in regional countries such as Nepal, Bangladesh, and Russia with the reported prevalence of 73%, 50%, and 36% respectively [26]. Similarly, UK has witnessed a rise in cases due to the delta variant during the first three weeks of June 2021 with health authorities fearing a possible third wave. The first case of B.1.617.2 from Pakistan was a male from Islamabad whose sample was collected in May 2021 and sent to NIH for laboratory testing. We found a high number of delta variant cases (n=8) from Islamabad followed by Azad Kashmir (n=1) and Rawalpindi (n=1) in the current study highlighting the hotspot regions of the country. To restrict these VOCs from community transmission, efficient screening and strict quarantine protocols for international flights need to be implemented.

For the first time in Pakistan, we have also found one significant mutation E484Q in a patient infected with B.1.617.2, that travelled back from Bahrain. The E484Q mutation has a prevalence of only 0.1% in B.1.617.2 as of June 29, 2021. The viruses with the mutations, L452R and E484Q mutations are more resistant to monoclonal antibodies, including bamlanivimab, and convalescent plasma [27]. Additionally, structural analysis of mutations in the furin cleavage site, L452R, E484Q, and P681R revealed increased ACE2 binding, increasing transmissibility.

Pakistan has confronted a four-month-long (March-June 2021) third wave of COVID-19 with the highest positivity rate reaching 11.6% in April and has declined since then to 2.35% as of 20 June 2021 [28]. Pakistan has recently eased the lockdown restrictions i.e., opening of restaurants, tourism sector, educational institutes, outdoor marriage ceremonies starting from May 24, 2021 [29]. However, the circulation of delta variant (the most transmissible form of the virus) in the indigenous population provides an early warning to national health authorities to take timely decisions. This also highlights the need for enhanced genomic surveillance to track the virus spread and devise suitable interventions.

## Data Availability

All the sequences generated in the current study are submitted to the GISAID

## DATA AVAILABILITY

All sequences generated in this study have been submitted to GISAID under the following accession numbers: EPI_ISL_2313081, EPI_ISL_2313082, EPI_ISL_2313084, EPI_ISL_2313086, EPI_ISL_2313098, EPI_ISL_2313099, EPI_ISL_2313112, EPI_ISL_2313114, EPI_ISL_2314185, EPI_ISL_2314809, EPI_ISL_2434174, EPI_ISL_2434247, EPI_ISL_2434781, EPI_ISL_2434972, EPI_ISL_2434976, EPI_ISL_2434982, EPI_ISL_2438547, EPI_ISL_2438598, EPI_ISL_2438599, EPI_ISL_2438665, EPI_ISL_2438666, EPI_ISL_2438683 We declare no competing interest.

